# Long-Term Daily Chlorhexidine Foot Cleansing Reduces Staphylococcal Burden on the Feet of People with Prior Diabetic Foot Complications

**DOI:** 10.64898/2026.05.14.26352248

**Authors:** Michelle R. Bode, Alison D. Lydecker, Gwen L. Robinson, Mary-Claire Roghmann, Lindsay R. Kalan

## Abstract

**Background:** Individuals with diabetes remain at high risk for diabetic foot ulcers (DFUs) even after ulcer healing. Dysbiosis of the skin microbiome has been associated with ulcer formation. Topical chlorhexidine gluconate (CHG) is a broad-spectrum antiseptic commonly used to reduce microbial burden. In a prior randomized clinical trial comparing daily CHG foot treatment with soap-and-water treatment, no statistically significant reduction in new DFUs was observed, prompting evaluation of whether CHG produced durable changes in the skin microbiota.

**Objective:** To compare changes in foot skin microbiota (including bacterial bioburden, diversity, and community composition) associated with daily CHG versus soap-and-water use over one year in people with diabetes and prior foot complications.

**Methods:** In a single-center, double-blind, placebo-controlled randomized trial, 87 participants were randomized to daily CHG wipes or soap-and-water wipes for 12 months. Foot swabs were collected at baseline, 3 and 12 months, and 4 weeks post-treatment. Bacterial bioburden was quantified. Microbiota composition was assessed using 16S rRNA and ITS amplicon sequencing.

**Key Results:** CHG treatment significantly reduced bacterial bioburden, increased microbial diversity, and altered community composition, including sustained reductions in *Staphylococcus* abundance. Several microbiota changes persisted more than 4 weeks after treatment cessation. Soap⍰and⍰water treatment showed similar but smaller and largely nonsignificant trends.

**Conclusions:** Daily CHG use durably modifies foot skin microbiota in high-risk individuals with diabetes. However, this alone may be insufficient to prevent new foot complications, highlighting the need for additional interventions. These findings have implications for long-term CHG use in populations at risk for staphylococcal infections.

**Key Points:** Long-term daily chlorhexidine foot cleansing in adults with diabetes reduces *Staphylococcus* abundance and overall bacterial burden more effectively than soap-and-water, with microbiota effects persisting for up to one month after treatment cessation, demonstrating durable pathogen suppression.

## Introduction

### Background

Diabetes affects more than 1 in 10 adults in the United States^1^ and is associated with serious complications, including diabetic foot ulcers (DFUs), which occur in approximately 19–34% of individuals during their lifetime.^2^ DFUs are a major cause of morbidity because more than 20% become infected and lead to lower extremity amputation, resulting in loss of mobility and independence.^3^ Once a DFU heals, patients remain at high risk for recurrence—particularly during the first year in which nearly 40% recur—because underlying pathophysiologic factors such as neuropathy and peripheral arterial disease (PAD) persist.^4^

Current prevention strategies emphasize daily foot inspection and hygiene^5^, yet recurrence rates remain unacceptably high. The role of the skin microbiota in DFU development and recurrence is hypothesized but not fully understood.^6^ The skin serves as a critical barrier and harbors a diverse microbial community that influences immunity and wound healing.^6^ Dysbiosis—imbalances in microbial composition—has been implicated in impaired wound healing and chronic inflammation.^7^ Prior work demonstrates that the feet of individuals with diabetes harbor an increased load of *Staphylococcus aureus* compared to those without diabetes^8^, and our unpublished work confirmed higher bacterial bioburden and lower microbial diversity in patients at high risk for DFU. These findings suggest that microbial composition may contribute to poor wound healing after minor trauma, a key initiating event in ulcer formation.^4^

### Rationale

Topical chlorhexidine gluconate is a broad-spectrum antiseptic widely used in healthcare settings to reduce microbial burden and prevent infections. Its application to outpatient foot care represents an innovative approach to DFU prevention. This led us to perform a single site randomized clinical trial.^9^ Daily use of CHG wipes for one year was compared with soap-and-water wipes in veterans with diabetes at high risk for foot complications. This intervention did not translate into a statistically significant reduction in new foot complications compared with soap-and-water (hazard ratio, 0.83; 95% CI, 0.39–1.80). However, the intervention was well tolerated, and adherence was high, underscoring the feasibility of structured foot hygiene as a preventive strategy.

One potential explanation for the lack of clinical benefit is that both study arms improved foot hygiene and produced similar microbiota shifts. Understanding these microbiota changes is critical for designing future interventions that target microbial signatures predictive of ulceration.^6^

### Objective

The objective of this study is to compare changes in skin microbiota—specifically composition and bioburden—between CHG wipes and soap-and-water wipes in individuals with diabetes and prior foot complications. We hypothesize that CHG use will significantly reduce microbial load and alter community structure compared with soap-and-water, providing insight into the long-term use of topical chlorhexidine and the findings of our clinical trial.

## Methods

### Study Design

We conducted the Preventing Diabetic Foot Ulcers Through Cleaner Feet trial as a single-center, double-blind (participant, outcome assessor), placebo-controlled, parallel group, phase 2b randomized clinical trial (RCT) at the Baltimore Veterans Affairs (VA) Medical Center from 2019 to 2023.^9^ Participants were randomly assigned, in a 1:1 ratio, to receive either wipes containing 2% chlorhexidine or wipes containing soap-and-water. Participants used one wipe daily for a year to clean their feet. Participants were ambulatory adult diabetics at high risk of a new DFU who did not have a current foot wound. Additionally, for this analysis, participants must have had a resolved prior foot complication (chronic foot ulcer, foot infection, or partial foot amputation) and must not have a current wound on their right foot to be included. An eSwab sample was collected from the pad of the right foot of each participant at the following timepoints: pre-intervention, 3 months on intervention, 12 months on intervention and 4 weeks post-intervention. All participants provided informed written consent. The study was approved by the University of Maryland Baltimore Institutional Review Board and the VA Research and Development Committee of the VA Maryland Health Care System (VAMHCS). The study is listed on ClinicalTrials.gov (NCT03503370).

The associations between treatment group and participant characteristics in Table 1 were measured using the χ^2^ test or Fisher’s exact test for categorical variables, and the Student’s t test or the Wilcoxon rank-sum test was used for continuous variables. All statistical tests were 2-tailed, and p-values < 0.05 were considered statistically significant. All statistical analyses were conducted using Stata 15 software (Stata Corp., College Station, TX).

**Table 1:** Demographic and Health Characteristics of Randomized Participants at Baseline by Treatment Group.

| Characteristic* | Chlorhexidine Group<br>(n=47) | Soap and Water Group<br>(n=40) | Total<br>(n=87) | P value† |
| --- | --- | --- | --- | --- |
| **Demographics:** |  |  |  |  |
| Age at enrollment (years), mean±SD | 67±7 | 65±8 | 6 ±8 | 0.31 |
| Male | 46 (98) | 39 (98) | 85 (98) | 1.00 |
| Race |  |  |  | 0.95 |
| American Indian or Alaskan Native | 0 (0) | 1 (3) | 1 (1) |  |
| Asian | 0 (0) | 0 (0) | 0 (0) |  |
| Black or African American | 30 (64) | 26 (65) | 56 (64) |  |
| Native Hawaiian or Other Pacific Islander | 1 (2) | 0 (0) | 1 (1) |  |
| White | 15 (32) | 13 (33) | 28 (32) |  |
| Prefer not to answer | 1 (2) | 0 (0) | 1 (1) |  |
| Ethnicity |  |  |  | 1.00 |
| Hispanic or Latino | (0) | 0 (0) | 0 (0) |  |
| Not Hispanic or Latino | 46 (98) | 40 (100) | 86 (99) |  |
| Prefer not to answer | 1 (2) | 0 (0) | 1 (1) |  |
| **Diabetes related factors:** |  |  |  |  |
| Most recent BMI, mean±SD | 32±6 | 30±5 | 31±6 | 0.12 |
| Most recent HbA1c, (%) mean±SD | 8.0±1.6 | 8.1±2.0 | 8.0±1.9 | 0.80 |
| Peripheral neuropathy | 33 (72) | 24 (60) | 57 (66) | 0.26 |
| Severe peripheral arterial disease by Arterial Brachial Index | 2 (4) | 5 (13) | 7 (8) | 0.24 |
| Most recent creatinine, median (IQR) (mg/dL) | 1.1 (1.0, 1.5) | 1.1 (0.9, 1.5) | 1.1 (0.9, 1.5) | 0.55 |
| Dialysis dependent | 3 (6) | 6 (15) | 9 (10) | 0.29 |
| **Foot Conditions:** |  |  |  |  |
| Months from healing of most recent healed foot complication to randomization, median (IQR) | 13 (5, 40) | 7 (3, 29) | 13 (4, 37) | 0.23 |
| *S. aureus* colonization on feet as determined by culture |  |  |  |  |
| Pre-treatment (n=84) | 9 (20) | 3 (8) | 12 (14) | 0.21 |
| After 3 months of treatment (n=75) | 8 (21) | 4 (12) | 12 (17) | 0.36 |
| Post treatment (n=68) | 6 (17) | 3 (10) | 9 (14) | 0.49 |
| **Co-morbidities:** |  |  |  |  |
| Hypertension | 42 (89) | 36 (90) | 78 (90) | 1.00 |
| Hyperlipidemia | 37 (79) | 34 (85) | 71 (82) | 0.45 |
| History of CVA or TIA | 4 (9) | 5 (13) | 9 (10) | 0.73 |
| Coronary Artery Disease | 13 (28) | 10 (25) | 23 (26) | 0.78 |
| Congestive heart failure | 7 (15) | 3 (8) | 10 (11) | 0.33 |
| Chronic venous insufficiency | 15 (32) | 8 (20) | 23 (26) | 0.21 |
| Currently smoke | 9 (19) | 6 (15) | 15 (17) | 0.61 |

\* Data are presented as number (percentage) of participants unless otherwise indicated.
† P values are from the $\chi^2$ test or Fisher's exact test for categorical variables, and the Student's t test or the Wilcoxon rank-sum test for continuous variables, as appropriate.

### Microbiota Analysis: 16S dPCR, 16S rRNA, ITS sequencing

Bacterial bioburden was assessed by quantifying a 500bp region in the V3–V4 region of the 16S rRNA gene using digital PCR (dPCR).^10^ The assay was run using 96-well 8.5k Nanoplates, 1µl of sample per 12 µl of reaction on a Qiagen QIAcuity eight. Qiagen’s standard cycling parameters were used. Analysis was performed using the R statistical programming language (v2025.09.0+387).^11^ Data were prepared and analyzed using the tidyverse package (v2.0.0).^12^ To compare bioburden between samples, the concentration measured in counts/µl was converted to a log10 scale. For each treatment, samples were separated by visit to track overall changes in bioburden. Significant differences from pre-treatment were determined using the Wilcoxon rank-sum test with Holm–Bonferroni p-value adjustment at a 95% confidence level.

Amplicon sequencing was performed on an Illumina MiSeq platform at the University of Maryland Institute for Genome Sciences. Raw V3-V4 amplicon 16S reads were initially processed using fastp (v0.24.0)^13^ to trim adapters and remove low-quality sequences. Changes in sequence quality were assessed with FastQC (v0.24.0)^14^ and visualized using MultiQC (v1.32).^15^ Additional quality filtering was performed using the QIIME2 (v2024.10)^16^ Amplicon Distribution workflow. DADA2^17^ was used to denoise, filter chimeras, truncate sequences (280 bp for forward reads and 240 bp for reverse reads), and merge reads. VSEARCH^18^ was then used to cluster reads into 99% operational taxonomic units (OTUs). OTUs were taxonomically classified using a Naive Bayes classifier implemented in scikit-learn, pre-trained on the SILVA full-length database (v138.21).^19^

Quality filtering of raw ITS1 reads followed a similar pipeline to 16S processing, with modifications to account for higher variability in sequence length. These included additional trimming of the poly-G tail in fastp, no length truncation, removal of short reads (<100 nts) with VSEARCH, and clustering into 97% OTUs. Taxonomic classification was performed using BLAST+ on the full UNITE+INSD dataset for Fungi (v19.02.20252).^20^

Eleven mock communities, 21 sample collection negative controls (4 from e-swabs and 17 from storage glycerol), and 8 PCR negative controls were also sequenced to ensure validity of results.

### Amplicon Sequencing

Final sample preparation and statistical analysis were performed using R. Data were imported, organized, and analyzed with the phyloseq package (v1.52.0).^21^ The metadata table was cleaned to ensure variables had correct names and data structures for downstream analysis. Unclassified top-abundant sequences were queried using Nucleotide BLAST against the rRNA/ITS database via the web service provided by the National Center for Biotechnology Information (NCBI) (https://blast.ncbi.nlm.nih.gov/Blast.cgi). Sequences were then reclassified to the taxonomic level where top hits reached consensus. OTUs with fewer than five reads were removed to reduce noise and filtered at the kingdom level to retain only sequences identified as Bacteria for 16S and Fungi for ITS.

The Decontam package (v1.28.0)^22^ was used to remove contaminants identified by high abundance in negative controls relative to samples. Rarefaction curves generated with the vegan package (v2.71)^23^ were used to determine minimum sample size cutoffs of 2,413 sequences for 16S and 701 sequences for ITS data. Data were rarefied to the minimum sample size for community analysis and processed in triplicate to ensure consistency. A non-rarefied version of the dataset was retained for relative abundance analysis. For all analyses, OTUs were aggregated to the genus level using tax_glom() to increase confidence in taxonomic identification accuracy.

Microbial community diversity was analyzed using phyloseq and visualized with ggplot2 and ggpubr. Alpha diversity measures, including observed species richness and the Shannon diversity index, were calculated using plot_richness(). For each treatment, samples were grouped by visit to observe changes in diversity over time. Significant differences between two groups were determined using the Wilcoxon rank-sum test with Holm–Bonferroni p-value adjustment at a 95% confidence level.

To analyze beta diversity, Bray–Curtis dissimilarity between samples were calculated. To visualize shifts in the microbiome, distances were ordinated and plotted using nonmetric multidimensional scaling (NMDS). Differences among multiple groups were assessed using PERMANOVA with adonis() in the vegan package.

To assess relative abundances of genera in each sample, samples were split by treatment group, and genera with fewer than 20 total reads were removed. To normalize sample size, sequence counts were converted to relative abundances. The sum of relative abundances across samples was used to select the top 15 genera in each treatment group, with remaining genera grouped as “Other”. The genus classified as *Methylobacterium-Methylorubrum* was renamed to its more commonly used name, *Methylobacterium*, for visualization. Significant differences from the pre-treatment group were determined using the Wilcoxon rank-sum test with Holm–Bonferroni p-value adjustment at a 95% confidence level. Correlations between bioburden and the relative abundance of *Staphylococcus* were assessed using Spearman rank correlation at a 95% confidence level.

### Statistical Analysis and Data Visualization

All statistical analysis and data visualization were done in the R statistical environment version v2025.09.0+387. Detailed code to generate each figure can be found in the data availability section.

## Results

### Study Design and Cohort Characteristics

Eighty-seven veterans with prior foot ulcers were enrolled in this study. The baseline demographics, clinical risk factors, and comorbidities of the 87 randomized participants were similar between treatment arms (Table 1). Fourteen percent of participants were colonized with *Staphylococcus aureus* on their feet immediately prior to starting the intervention, and there was no statistically significant difference by treatment arm at any timepoint. Throughout the study, samples were collected from each patient, with some limitations in adherence due to COVID19 restrictions (Figure 1). Pretreatment swab samples were collected from all patients; after processing and quality checking, 84 were sequenced. Participants were then randomized into daily foot washing groups using either chlorhexidine (n=47) or soap-and-water (n=40), with subsequent samples collected after 3 months of treatment (chlorhexidine n=38; soap-and-water n=33), 12 months of treatment (chlorhexidine n=31; soap-and-water n=28), and approximately four weeks after treatment completion (chlorhexidine n=36; soap-and-water n=31).

**Figure 1:**
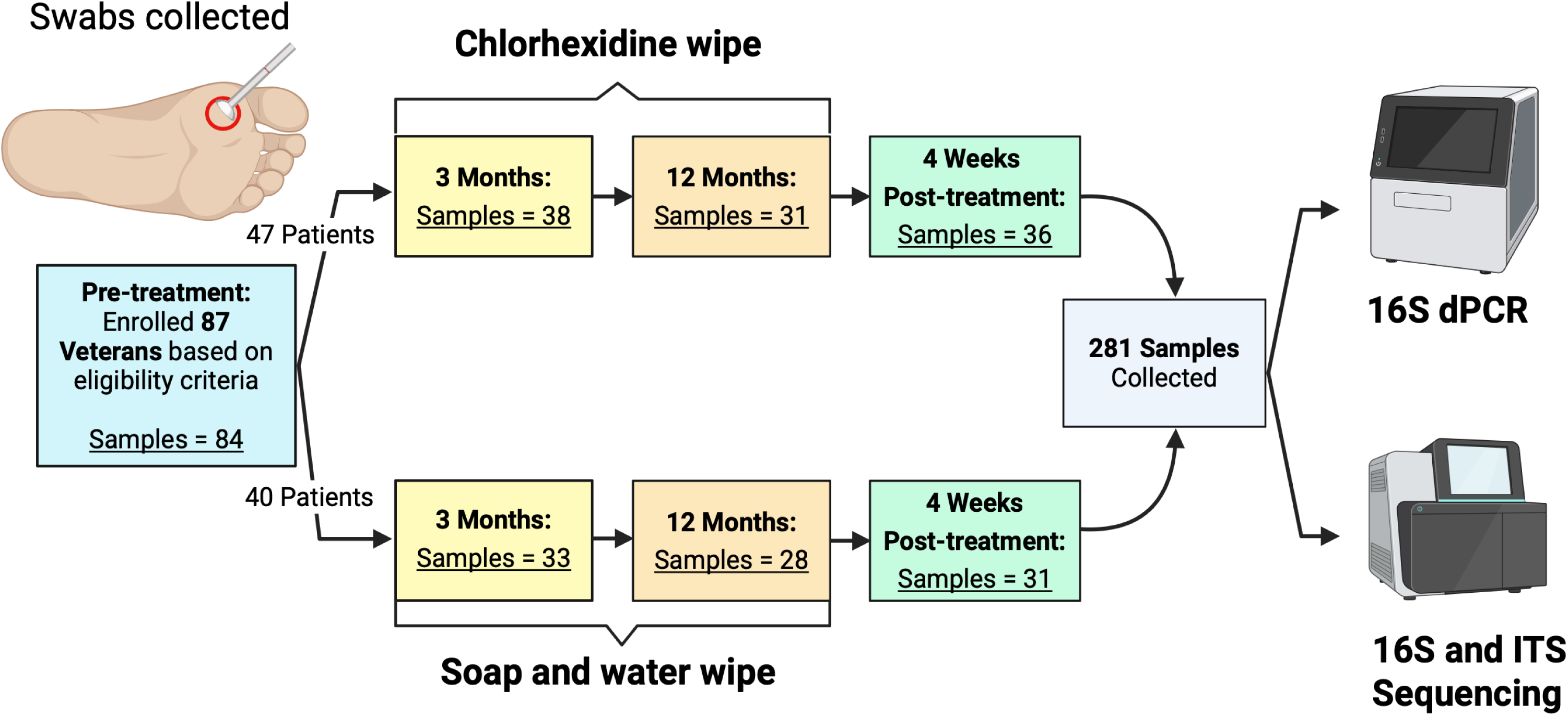
Overview of study designed to observe the effects that daily chlorhexidine foot washing for one year has on the microbiome. After pre-screening, 87 veterans with diabetes who had previous foot complications were enrolled in this study. The microbiome for each patient over time was assessed from eSwab samples collected from the pad of their right foot. Missing samples from follow-up visits were due to COVID-19 restrictions preventing attendance. Prior to treatment, 84 swab samples were collected. Patients were then blindly split into treatment groups, where they were given either chlorhexidine (n = 47) or soap-and-water (n = 40) wipes for daily foot washing. Samples were collected from patients after 3 months (chlorhexidine n = 38, soap-and-water n = 33) and 12 months (chlorhexidine n = 31, soap-and-water n = 28) of treatment. Treatment was completed after one year, but additional samples were collected from patients approximately 4 weeks post-treatment (chlorhexidine n = 36, soap-and-water n = 31). Swab samples were then sequenced for the fungal ITS1 region and the bacterial 16S region for taxonomic identification and relative abundances of the microbes present. Total 16S was also quantified using dPCR to assess bacterial bioburden. Created with BioRender. [Kalan, L. (2026) https://BioRender.com/8kouyw5].

The foot skin microbiome was measured via amplicon sequencing of the V3-V4 region of the bacterial 16S rRNA gene and the ITS1 region of the fungal rRNA cistron. For 16S analysis, a total of 219 samples (78%) were retained after quality control filtering and implementing minimum sample size cutoffs. Of the total 16S samples maintained, 122 (56%) were chlorhexidine treated (pre-trt n=42, 3 months n= 27, 12 months n=25, post-trt n=28) and 97 (44%) were soap-and-water treated (pre-trt n=27, 3 months n=24, 12 months n=22, post-trt n=24). In total, 3,714 16S OTUs were identified, with an average of 529 (95% CI = 227-831) counts per sample. After rarefaction, 3,605 OTUs remained, with a standard deviation of 6 OTUs between replicates created to ensure consistent results. For ITS analysis, a total of 173 samples (62%) were retained after quality control filtering and implementing minimum sample size cutoffs. Of the total ITS samples maintained, 96 (55%) were chlorhexidine treated (pre-trt n=26, 3 months n= 25, 12 months n=17, post-trt n=28) and 77 (45%) were soap-and-water treated (pre-trt n=26, 3 months n=20, 12 months n=14, post-trt n=17). In total, 263 ITS OTUs were identified, with an average of 4,352 (95% CI: 2346-6358) counts per sample. After rarefaction, 257 OTUs remained, with no standard deviation between replicates created to ensure consistent results.

### CHG Application Does Not Have a Significant Effect on Fungal Communities

No significant shifts in the presence of fungal genera were detected over the course of chlorhexidine and soap-and-water treatment (Supplemental Figure 1A). Bray-Curtis dissimilarity from pre-treatment samples remained consistently high for both treatments with no variation across time points (Supplemental Figure 1B). Dissimilarity between chlorhexidine samples ranged from 0.81 to 0.89 (95% CI) and 0.76 to 0.88 for soap-and-water samples (95% CI). No significant differences in fungal presence were seen between samples with active treatment (samples collected after 3 and 12 months of treatment) and no active treatment (samples from pre-treatment and post-treatment) (Supplemental Figure 1C). Fungal presence differed widely across all samples with no distinction between treatments (Supplemental Figure 2A).

*Malassezia* was the most prevalent genus overall, followed by *Saccharomyces*, *Trichophyton*, and *Candida*. Average Shannon diversity remained consistent over the course of both treatments, with no significant changes from pre-treatment samples (Supplemental Figure 2B). For chlorhexidine treatment, the lowest average Shannon diversity was 0.78 (95% CI = 0.54-1.02) at 3 months and the highest was 0.89 (95% CI = 0.66-1.12) at pre-treatment. For soap-and-water treatment, the lowest average Shannon diversity was 0.83 (95% CI = 0.69-0.99) at 3 months and the highest was 0.89 (95% CI = 0.71-1.05) at post-treatment. Due to the limited detection of the observed effect of chlorhexidine application on the fungal microbiome, the remainder of the presented analysis focuses exclusively on the bacterial microbiome.

### Microbial Bioburden and Intra-Sample Diversity Shifts After CHG Application

Bacterial bioburden was assessed using digital PCR (dPCR) at all visits. After removing samples with no reads, a total of 277 samples (99%) were maintained. Of the total samples, 148 (53%) were chlorhexidine treated (pre-trt n=45, 3 months n=38, 12 months n=31, post-trt n=34) and 127 (47%) were soap-and-water treated (pre-trt n=38, 3 months n=32, 12 months n=28, post-trt n=31). Chlorhexidine treatment resulted in a significant reduction of bioburden compared to pre-treatment (Wilcoxon rank-sum test; 3 months p <0.0001, 12 months p<0.01; Supplemental Figure 3A; Figure 3A). Prior to treatment, samples had an average 16S log_10_ concentration of 2.55 (95% CI = 2.26-2.84) that substantially decreased after 3 months of treatment to 1.78 (95% CI = 1.44-2.06). This reduction in bioburden was seen for 72% of patients. After 12 months, average bioburden increased slightly to 1.94 (95% CI = 1.54-2.34). This increase in bioburden from 3 months of treatment was seen for 55% of individuals. Following treatment completion, average bioburden increased substantially to 2.44 (95% CI = 2.05-2.84) and was no longer significant from samples prior to treatment. This increase in bioburden from 12 months of treatment was observed in 74% of patients.

Although bioburden decreased with soap-and-water treatment, the differences from pre-treatment samples were not significant, largely due to high inter-patient variability. (Supplemental Figure 3A; Figure 2A). Prior to treatment, samples had an average 16S log_10_ concentration of 2.16 (95% CI = 1.88-2.45) that decreased after 3 months of treatment to 1.91 (95% CI = 1.56-2.26). This reduction in bioburden was seen for 57% of patients. After 12 months, average bioburden increased marginally to 1.89 (95% CI = 1.54-2.25). This increase in bioburden from 3 months of treatment was observed in 56% of individuals. Following treatment completion, average bioburden increased substantially to 2.39 (95% CI = 2.04-2.75) and was no longer significant from samples prior to treatment. This increase in bioburden from 12 months of treatment was observed in 68% of patients.

**Figure 2:**
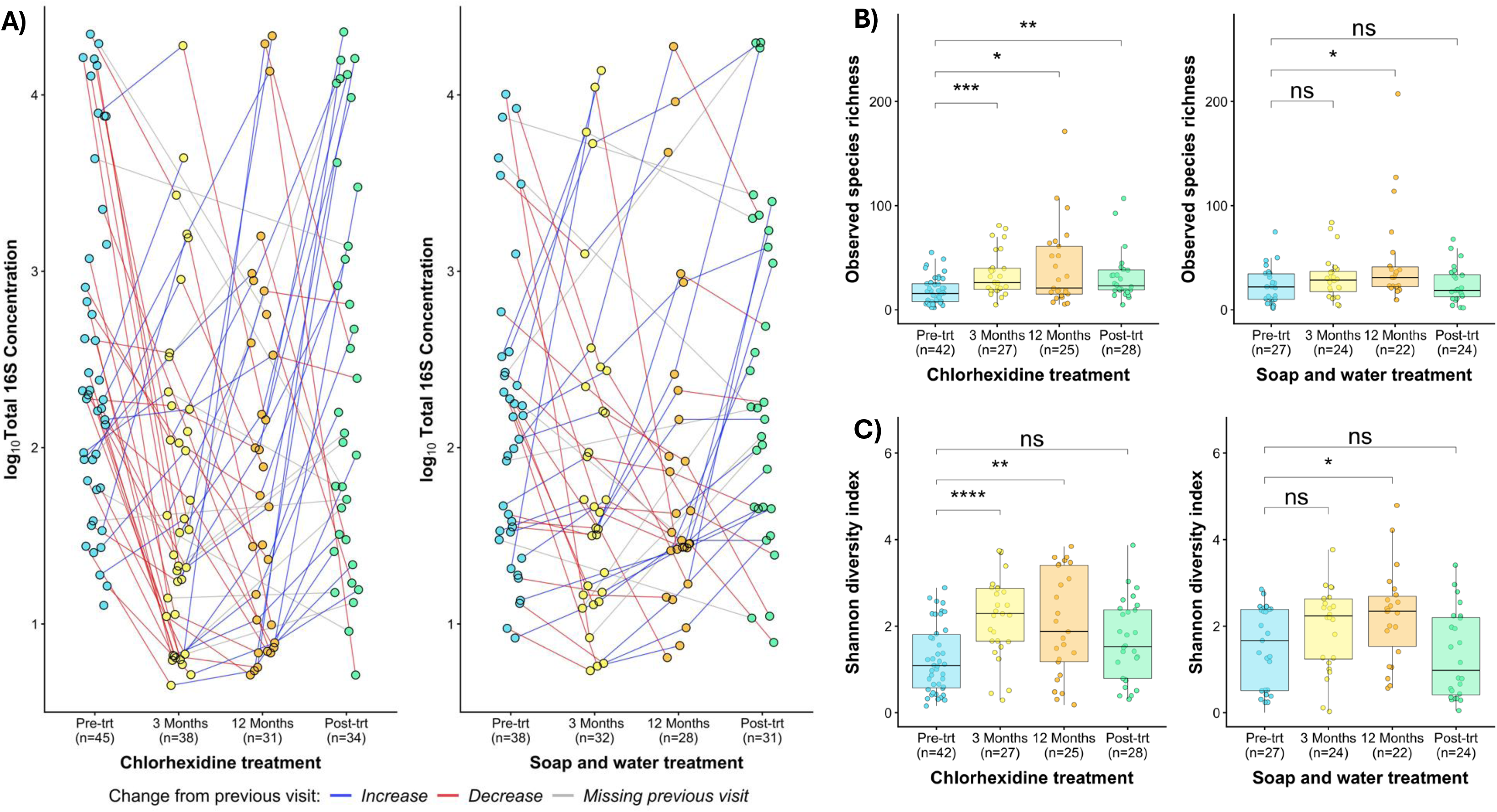
Chlorhexidine treatment results in a decrease in total bacterial bioburden and an increase in Shannon diversity and observed richness. (A) The log concentrations of total 16S were measured using dPCR and compared separately over the course of treatment for chlorhexidine (pre-trt n = 45, 3 months n = 38, 12 months n = 31, post-trt n = 34) and soap-and-water (pre-trt n = 38, 3 months n = 32, 12 months n = 21, post-trt n = 31). Changes at an individual level over consecutive visits were assessed by connecting lines between samples from the same patient. The color of the lines represents an increase from the prior visit (blue), a decrease from the prior visit (red), and cases where no prior visit was present (grey). (B) For bacterial diversity analysis, samples with low counts were removed, OTU counts were normalized with rarefaction, and data were merged to the genus level. Observed species richness of the OTUs in each sample was measured and compared separately over the course of treatment with chlorhexidine (pre-trt n = 42, 3 months n = 27, 12 months n = 25, post-trt n = 28) and soap-and-water (pre-trt n = 27, 3 months n = 24, 12 months n = 22, post-trt n = 24). (C) Shannon diversity was measured for the same groups described in Figure 2B. Statistically significant differences in averages between pre-treatment samples and subsequent visits were calculated using a Wilcoxon rank-sum test, with Benjamini–Hochberg p-value adjustment. * p<0.05, ** p<0.01, *** p<0.001, **** p<0.0001.

We then assessed genus level changes in overall bacterial diversity of each sample overtime using the 16S amplicon data. Chlorhexidine treatment resulted in a significant increase in observed species richness compared to pre-treatment (Wilcoxon rank-sum: 3 months p < 0.001, 12 months p < 0.05, post-treatment p < 0.01; Figure 2B). The average number of OTUs per sample increased after treatment from 18 (95% CI = 14-22) OTUs to 34 (95% CI = 26-43) after 3 months and 41 (95% CI = 25-57) OTUs after 12 months. At the completion of treatment, average richness decreased to 31 (95% CI = 22-40) OTUs per sample. Soap-and-water application resulted in a similar trend in average richness that only reached significance after 12 months (p < 0.05) (Figure 2B). Chlorhexidine treatment also resulted in a significant increase in Shannon diversity compared to pre-treatment (Wilcoxon rank-sum: 3 months p < 0.0001, 12 months p < 0.01; Figure 2C). Once treatment stopped, the average diversity index decreased and was no longer significantly different from pre-treatment. Soap-and-water application again resulted in a similar trend, but significant differences were not observed until after 12 months (p < 0.05) (Figure 2C).

### Microbial Community Composition Shifts After CHG Treatment

To assess changes in the composition of the core microbiome for each treatment and over time, we calculated Bray–Curtis dissimilarity metrics between samples and time points (Figure 3; Supplemental Figure 3B&C). Chlorhexidine application resulted in statistically significant clustering of samples by time point (PERMANOVA: p < 0.001, R² = 0.090; Figure 3A; Supplemental Figure 3B). After treatment ended, microbial community composition shifted back to a pre-treatment state. Samples collected at different time points clustered by whether they had active treatment (samples collected after 3 and 12 months of treatment) or no active treatment (samples from pre-treatment and post-treatment) (p < 0.001, R² = 0.090) (Figure 3B). At the individual level (n=21), similar directional shifts were seen after 3 and 12 months of treatment indicating a consistent change in the microbiome across samples (p < 0.01, R² = 0.11) (Figure 3C; Supplemental Figure 3C).

**Figure 3:**
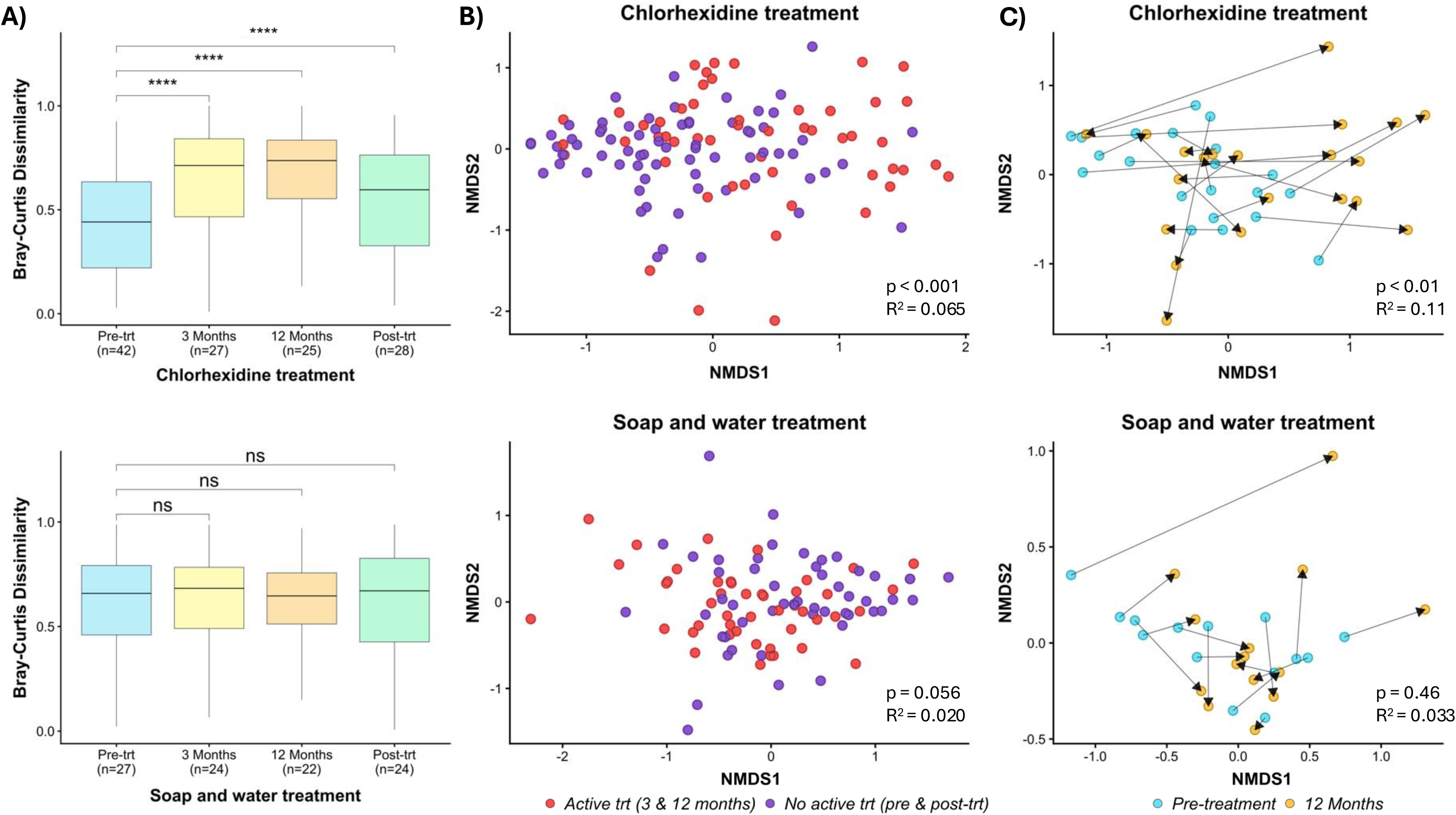
Chlorhexidine treatment causes a significant and consistent directional shift in the core microbiome. For bacterial diversity analysis, samples with low counts were removed, OTU counts were normalized with rarefaction, and data were merged to the genus level. Samples were then separated by visit for both chlorhexidine (pre-trt n = 42, 3 months n = 27, 12 months n = 25, post-trt n = 28) and soap-and-water (pre-trt n = 27, 3 months n = 24, 12 months n = 22, post-trt n = 24) treatments. (A) Differences in the relative abundances of genera in each sample were measured using Bray–Curtis dissimilarity. The average distance of all samples from pre-treatment samples was measured and separated by visit to observe whether distance increased with treatment. Statistically significant differences between groups for pairwise comparisons were calculated using a Wilcoxon rank-sum test, with Benjamini–Hochberg p-value adjustment. * p < 0.05, ** p < 0.01, *** p < 0.001, **** p < 0.0001. (B) Bray–Curtis distances between samples were visualized on an NMDS plot. Samples were grouped by whether there was active treatment (3-month and 12-month) shown in red, or no active treatment (pre-treatment and post-treatment) shown in purple. (C) To observe changes at the individual level, Bray–Curtis distances between samples from pre-treatment and after 12 months of treatment were visualized on an NMDS plot, with lines connecting samples from the same patient. Arrows indicate the direction of shift from pre-treatment to after 12 months of treatment for each individual. Statistically significant differences between groups were calculated using PERMANOVA, with Benjamini–Hochberg p-value adjustment.

No significant separation of samples over time was observed in the soap-and-water treatment group (Figure 3A and Supplemental Figure 3B). Albeit not statistically significant, there was a trend where weak clustering appeared based on active treatment time (samples collected after 3 and 12 months of treatment) or no active treatment (samples from pre-treatment and post-treatment) (p=0.056, R^2^ = 0.020) (Figure 3B). At the individual level (n=14), random directional shifts were observed after 3 and 12 months of treatment indicating inconsistent changes in the microbiome across samples (Figure 3C and Supplemental Figure 3C).

### CHG Application Results in a Reduction of Staphylococcus Abundance

To better understand the changes in microbiome composition that occurred in response to chlorhexidine or soap-and-water application, the relative distribution of the top 15 genera for each treatment was examined over timepoints (Figure 4A). Similar genera were observed in both groups, with a higher proportion of Gram-positive compared to Gram-negative bacteria. *Staphylococcus* was the dominant genus, followed by *Corynebacterium*, *Enhydrobacter*, and *Kocuria*. With chlorhexidine treatment, the relative abundance of *Staphylococcus* was significantly reduced over time (Wilcoxon rank-sum: 3 months p <0.0001, 12 months p<0.0001, post-treatment<0.05; Supplemental Figure 4A; Figure 4B). Prior to treatment, samples had an average *Staphylococcus* relative abundance of 63% (95% CI = 55%-71%). After 3 months of treatment, 83% of patients showed an average 30% (95% CI = 18%-42%) decrease in *Staphylococcus* abundance. After 12 months, 75% of patients showed a slight recovery in *Staphylococcus* relative abundance. Upon completion of treatment, a substantial recovery in *Staphylococcus* abundance was observed for 75% of individuals, with the total average proportional abundance increasing to 47% (95% CI = 6%-34%).

**Figure 4:**
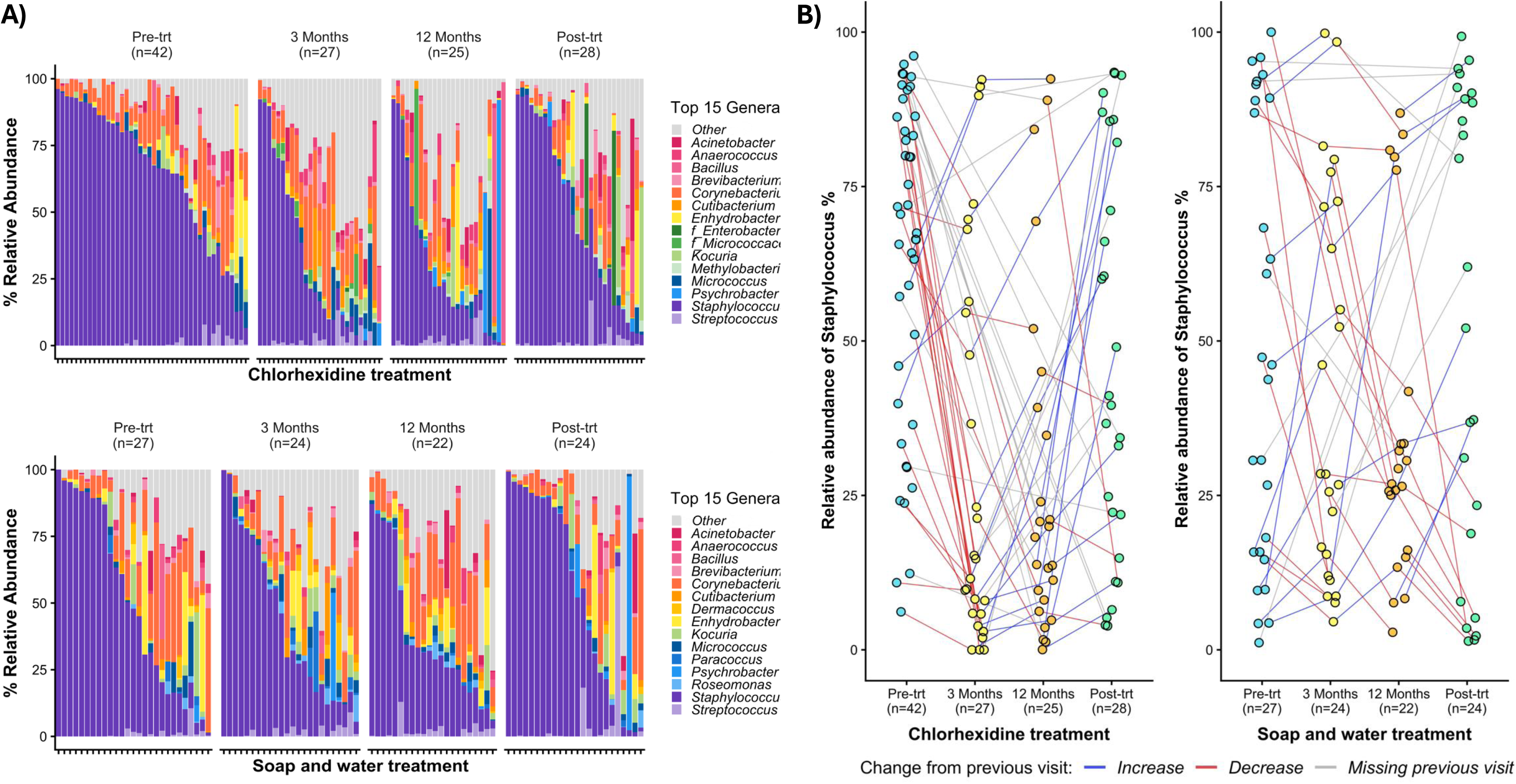
Chlorhexidine treatment causes a significant decrease in *Staphylococcus* relative abundance. (A) The relative abundances of the top 15 bacterial genera were compared separately over the course of treatment with chlorhexidine (pre-trt n = 42, 3 months n = 27, 12 months n = 25, post-trt n = 28) and soap-and-water (pre-trt n = 27, 3 months n = 24, 12 months n = 22, post-trt n = 24). (B) Changes in Staphylococcus abundance over consecutive visits were assessed at the individual level by connecting lines between samples from the same patient. The color of the lines represents an increase from the prior visit (blue), a decrease from the prior visit (red), and cases where no prior visit was present (grey).

In the soap-and-water treatment group, non-significant decreases in *Staphylococcus* abundance were observed over time (Supplemental Figure 4A; Figure 4B). Prior to treatment, samples had an average *Staphylococcus* relative abundance of 50% (95% CI = 36%-64%). After 3 months of treatment, 52% of patients showed an average decrease in *Staphylococcus* abundance, albeit to a much smaller extent than the chlorhexidine group, which had an average 3 month abundance of 42% (95% CI = 29%-56%). After 12 months, 69% of patients showed a decrease in *Staphylococcus* relative abundance. Upon completion of treatment, *Staphylococcus* abundance recovered back to an average of 53% (95% CI = 37%-69%). To determine if the overall absolute decrease in bacterial abundances measured via dPCR were proportional to *Staphylococcus* relative abundance, Spearman’s rank correlations were calculated (Supplemental Figure 4B). A significant correlation was found with both the application of chlorhexidine (R = 0.49, p <0.0001) and soap-and-water (R = 0.57, p <0.0001) suggesting the overall decrease in bacterial bioburden is driven by a decrease in *Staphylococcus* presence.

## Discussion

### Summary of results

Daily chlorhexidine treatment produced clear and consistent shifts in the foot skin microbiota over time that were greater than soap-and-water treatment. Compared with baseline, chlorhexidine reduced bacterial bioburden, increased overall biodiversity, and altered microbiota composition, most notably through a sustained decrease in *Staphylococcus* abundance. Similar patterns were observed with soap-and-water treatment, but these changes were smaller in magnitude. Importantly, more than four weeks after treatment cessation, participants in the chlorhexidine group continued to demonstrate lower bacterial bioburden, higher observed species richness, and lower relative *Staphylococcus* abundance compared to their own baseline samples. This indicates the effects of chlorhexidine treatment persisted beyond the active treatment period. Despite these measurable and durable microbiota changes, they did not translate into reduced ulcer recurrence, suggesting that lowering bacterial bioburden, particularly reducing *Staphylococcus,* is insufficient on its own to prevent diabetic foot ulcer recurrence.

### Comparison to Prior Studies

Most of the top genera identified concur with findings in previous studies.^24,25^ An unusual finding in our study was the high prevalence of *Enhydrobacter*. This taxon has been shown to be associated with older populations in other skin sites,^26^ consistent with the higher average age of participants in this study. The reduction in bioburden and the decrease in *Staphylococcus* align with our previous work examining the effects of chlorhexidine pre-surgical prep on the normal skin microbiome.^27–29^ This study and others reported changes in *Bacillus* and *Enterococcus* population with chlorhexidine treatment, which we did not observe due to a smaller baseline population of these taxa on the feet.^27,30,31^ Previous studies reporting no significant changes in the skin microbiome after chlorhexidine exposure are likely due to multiple factors. These include short exposure times and the inability to separate viable from non-viable organisms due to the ability of chlorhexidine to bind DNA from recently lysed cells. Indeed, Townsend *et al*. used a special viability PCR method to demonstrate large shifts in both bioburden and microbiome composition upon chlorhexidine skin bathing. In this study, the longitudinal sampling design allowed detection of chlorhexidine residual efficacy by sampling after chlorhexidine is washed from the skin. In both studies, the skin microbiome begins a return to baseline upon cessation of chlorhexidine application. This also highlights the role of long-term application for substantial effects.^32–34^ However, reduced effectiveness of treatment as indicated by rising microbial diversity over time may reflect the development of chlorhexidine resistance, which has been reported in prior studies.^27,35–39^ An alternative hypothesis is that that chlorhexidine as a broad spectrum antiseptic has its major effect on *Staphylococcus,* a dominant taxa on the skin.

The top fungal genera identified aligned with prior findings, including high variability between patients.^24,40^ Our study was not the first to observe marginal effects of chlorhexidine on fungi. Nascimento *et al*. performed a culture based study to observe if CHG treatment reduces *Candida* on skin and also observed no significant changes.^41^ To our knowledge no prior studies of the human skin microbiome have shown the effectiveness of CHG on reducing *Candida* populations but it has been reported in oral microbiome and *in vitro*.^42,43^

### Strengths and Limitations

This study has several important strengths. To our knowledge, it represents the largest randomized clinical trial to evaluate foot hygiene interventions in individuals with diabetes and prior foot complications, a population at particularly high risk for ulcer recurrence. The randomized, blinded design allowed direct comparison of daily chlorhexidine treatment with an optimized soap-and-water regimen, strengthening causal inference regarding microbiota effects. Longitudinal follow-up over 12 months, with additional post-treatment sampling, enabled assessment of both short-term and durable changes in the foot skin microbiota. In addition, the study employed a robust and comprehensive microbiota analytic approach, combining digital PCR with bacterial and fungal amplicon sequencing and rigorous quality control, providing high-resolution insights into microbial bioburden, diversity, and community composition.

Several limitations should also be considered. The study was conducted at a single center within the Veterans Affairs healthcare system, and the cohort was predominantly male and Black/African American. This limits generalizability to broader populations, including women and non-veterans and those outside the geographic area. Missing swab samples at some time points, largely attributable to disruptions related to the COVID-19 pandemic, reduced sample availability for some longitudinal analyses, and may have limited power to detect smaller microbiota changes over time. Finally, the use of 16S and ITS sequencing for taxonomic classification limits the ability to obtain accurate species level identification.

### Implications for Clinical Practice

Topical chlorhexidine has multiple established uses for preventing healthcare-associated infections, including daily bathing, preoperative skin antisepsis, catheter site care, and decolonization protocols for multidrug-resistant organisms. Our use is most like daily bathing, which is performed in intensive care units (ICUs) to prevent central line associate bloodstream infections and reduce the acquisition of MDROs. A meta-analysis found chlorhexidine bathing reduced hospital-acquired bloodstream infections by 41% (IRR 0.59, 95% CI 0.52-0.68) in ICUs.^44^ Chlorhexidine bathing is also used long term outside the hospital; for example, individuals who have undergone bone marrow transplantation and require prolonged intravenous access often use daily topical chlorhexidine to reduce bloodstream infection risk.^45^ Our findings demonstrate a sustained reduction in bacterial bioburden—driven predominantly by decreases in *Staphylococcus*—providing a potential mechanistic explanation for the protective effects of chlorhexidine against line associated infections. Notably, the persistence of microbiota changes for up to one month after discontinuation suggests that less-than-daily chlorhexidine use may still confer benefit. This has practical implications, as adherence to daily chlorhexidine bathing is frequently lower than expected due to patient reported barriers, including transient skin stickiness, feeling cold after application, and difficulty applying the product to the entire body.^46^

### Future Outlook and Conclusions

Here we demonstrate that daily foot treatment with chlorhexidine can significantly alter the skin microbiota and reduce infection associated pathogens, specifically *Staphylococci* compared to soap-and-water. While new foot complications between the two treatment groups did not differ, the overall prevalence was low. Future studies focusing on microbial markers to predict risk of ulcer recurrence will require a larger cohort for statistical power, including those at higher risk such as individuals with severe PAD, which were excluded in this study. However, our finding that chlorhexidine intervention can reduce *Staphylococcal* burden on the skin in a sustained manner suggests there may be a clinical benefit for populations at risk of *Staphylococcal* infection. This includes reducing infection risk even if ulceration occurs, as most diabetic foot ulcers are colonized by diverse *Staphylococcal* species with specific strains of *S. aureus* being associated with more severe outcomes.^47–49^ In summary, while preventing ulcer recurrence requires multi-faceted approaches including microbiological, physiological, and nutritional factors, targeted microbiome manipulation may serve to modulate long-term infection and severe outcome risks.

## Acknowledgements

The authors thank Maryland Genomics at the University of Maryland Institute for Genome Sciences for performing the amplicon sequencing.

## Contributions

M.R. and L.K. acquired funding, administered the project and provided resources and supervision. M.R. conceptualized the study. M.B. and A.L curated the data and performed the formal analysis. M.B. performed the investigation, validation and visualization of the data. M.R., L.K., M.B, G.R., and A.L contributed to the methodology, writing the first draft, and the review and editing of the final draft.

## Data availability statement

Sequence reads for this project can be found under NCBI BioProject PRJNA1457219. Code for analysis and generation of figures can be found on GitHub at: https://github.com/Kalan-Lab/CHG-Effect-on-the-Skin-Microbiome-of-People-with-Diabetes

---

**Supplemental Table 1: Self-reported Foot Care of Randomized Participants at Baseline by Treatment Group**

**Supplemental Figure 1: Chlorhexidine treatment caused no significant changes in the fungal microbiome.** For fungal diversity analysis, samples with low counts were removed, OTU counts were normalized with rarefaction, and data were merged to the genus level. Samples were then separated by visit for both chlorhexidine (pre-trt n = 26, 3 months n = 25, 12 months n = 17, post-trt n = 28) and soap-and-water (pre-trt n = 26, 3 months n = 20, 12 months n = 14, post-trt n = 17) treatments. (A) Differences in the relative abundances of genera in each sample were measured using Bray–Curtis dissimilarity and visualized on an NMDS plot. (B) The average distance of all samples from pre-treatment samples was measured and separated by visit to observe whether distance increased with treatment. (C) Bray–Curtis distances between samples were visualized on an NMDS plot. Samples were grouped by whether there was active treatment (3-month and 12-month) shown in red, or no active treatment (pre-treatment and post-treatment) shown in purple. Statistically significant differences between groups were calculated using PERMANOVA, with Benjamini–Hochberg p-value adjustment. Statistically significant differences between groups for pairwise comparisons were calculated using a Wilcoxon rank-sum test, with Benjamini–Hochberg p-value adjustment. * p < 0.05, ** p < 0.01, *** p < 0.001, **** p < 0.0001.

**Supplemental Figure 2: Chlorhexidine treatment does not cause significant changes in the relative abundance and Shannon diversity of fungal genera.** (A) The relative abundances of the top 15 fungal genera were compared separately over the course of treatment with chlorhexidine (pre-trt n = 26, 3 months n = 25, 12 months n = 17, post-trt n = 28) and soap-and-water (pre-trt n = 26, 3 months n = 20, 12 months n = 14, post-trt n = 17). (B) For fungal diversity analysis, samples with low counts were removed, OTU counts were normalized with rarefaction, and data were merged to the genus level. Shannon diversity was measured in each sample and compared separately over the course of each treatment. Statistically significant differences in averages between pre-treatment samples and subsequent visits were calculated using a Wilcoxon rank-sum test, with Benjamini–Hochberg p-value adjustment. * p < 0.05, ** p < 0.01, *** p < 0.001, **** p < 0.0001.

**Supplemental Figure 3: Chlorhexidine treatment causes significant decreases in bioburden and consistent directional shift in the core microbiome.** (A)The log concentrations of total 16S in each sample were measured using dPCR and compared separately over the course of treatment for chlorohexidine (pre-trt n= 45 months n =38, 12 months n=31, post-trt n=34) and soap-and-water (pre-trt n= 38 months n =32, 12 months n=21, post-trt n=31). (B) For bacterial diversity analysis, samples with few counts were removed, OTU counts were normalized with rarefaction and merged to genus level. Samples were then separated by visit for both chlorohexidine (pre-trt n= 42 months n =27, 12 months n=25, post-trt n=28) and soap-and-water (pre-trt n= 27, months n=24, 12 months n=22, post-trt n=24) treatments. The differences in the relative abundances of genera in each sample were measured using Bray-Curtis dissimilarity and visualized on an NMDS plot. (C) To observe changes in an individual, Bray-Curtis distances between samples from pre-treatment and after 12 months of treatment were visualized on an NMDS plot lines connecting samples from the same patient. Arrows indicate the direction of shift from pre-treatment to after 12 months of treatment for an individual. Statically significant differences between groups were calculated using PERMANOVA, with Benjamini-Hochberg p-value adjustment. Statically significant differences between groups for pairwise comparisons were calculated using a Wilcoxon-rank sum test, with Benjamini-Hochberg p-value adjustment. * p<0.05, ** p<0.01, *** p<0.001, **** p<0.0001.

**Supplemental Figure 4: Chlorhexidine treatment causes significant decreases in staphylococcus abundance that is correlated with a decrease in bioburden.** (A) The relative abundances of Staphylococcus in each sample, determined with 16S sequencing, were compared separately over the course of treatment with chlorohexidine (pre-trt n= 26 months n =25, 12 months n=17, post-trt n=28) and soap-and-water (pre-trt n= 26, months n=20, 12 months n=14, post-trt n=17). Statically significant differences in averages between pre-treatment samples and the following visits were calculated using a Wilcoxon-rank sum test, with Benjamini-Hochberg p-value adjustment. * p<0.05, ** p<0.01, *** p<0.001, **** p<0.0001. (B) The log concentrations of total 16S were measured using dPCR and compared to the relative abundances of Staphylococcus to observe if changes were proportional to each other. A significant linear correlation was measured using Spearman rank correlations.

## References

1. Gwira J, Fryar C, Gu Q. Prevalence of Total, Diagnosed, and Undiagnosed Diabetes in Adults: United States, August 2021–August 2023. NCHS Data Brief No. 516. Published online November 2024. doi:CS354814

2. Armstrong DG, Tan TW, Boulton AJM, Bus SA. Diabetic Foot Ulcers: A Review. JAMA. 2023;330(1):62–75. doi:10.1001/jama.2023.10578

3. McDermott K, Fang M, Boulton AJM, Selvin E, Hicks CW. Etiology, Epidemiology, and Disparities in the Burden of Diabetic Foot Ulcers. Diabetes Care. 2023;46(1):209–221. doi:10.2337/dci22-0043

4. Pecoraro RE, Reiber GE, Burgess EM. Pathways to diabetic limb amputation. Basis for prevention. Diabetes Care. 1990;13(5):513–521.

5. American Diabetes Association Professional Practice Committee for Diabetes*. 12. Retinopathy, Neuropathy, and Foot Care: Standards of Care in Diabetes-2026. Diabetes Care. 2026;49(Supplement_1):S261–S276. doi:10.2337/dc26-S012

6. Grice EA, Kong HH, Renaud G, et al. A diversity profile of the human skin microbiota. Genome Res. 2008;18(7):1043–1050. doi:10.1101/gr.075549.107

7. Tomic-Canic M, Burgess JL, O’Neill KE, Strbo N, Pastar I. Skin Microbiota and its Interplay with Wound Healing. Am J Clin Dermatol. 2020;21(Suppl 1):36–43. doi:10.1007/s40257-020-00536-w

8. Redel H, Gao Z, Li H, et al. Quantitation and composition of cutaneous microbiota in diabetic and nondiabetic men. JInfectDis. 2013;207(7):1105–1114. doi:10.1093/infdis/jit005;

9. Lydecker AD, Kim JJ, Robinson GL, et al. Chlorhexidine vs Routine Foot Washing to Prevent Diabetic Foot Ulcers: A Randomized Clinical Trial. JAMA Netw Open. 2025;8(2):e2460087. doi:10.1001/jamanetworkopen.2024.60087

10. Liu CM, Aziz M, Kachur S, et al. BactQuant: an enhanced broad-coverage bacterial quantitative real-time PCR assay. BMC Microbiol. 2012;12:56. doi:10.1186/1471-2180-12-56

11. The R Core Team. R: A Language and Environment for Statistical Computing, Version 4.5.3. Published online March 11, 2026. https://cran.r-project.org/doc/manuals/r-release/fullrefman.pdf

12. Wickham H, Averick M, Bryan J, et al. Welcome to the Tidyverse. The Journal of Open Source Software. 2019;4(43).

13. Chen S, Zhou Y, Chen Y, Gu J. fastp: an ultra-fast all-in-one FASTQ preprocessor. Bioinformatics. 2018;34(17):i884–i890. doi:10.1093/bioinformatics/bty560

14. FastQC A Quality Control tool for High Throughput Sequence Data. Accessed March 26, 2026. https://www.bioinformatics.babraham.ac.uk/projects/fastqc/

15. Ewels P, Magnusson M, Lundin S, Käller M. MultiQC: summarize analysis results for multiple tools and samples in a single report. Bioinformatics. 2016;32(19):3047–3048. doi:10.1093/bioinformatics/btw354

16. Bolyen E, Rideout JR, Dillon MR, et al. Reproducible, interactive, scalable and extensible microbiome data science using QIIME 2. Nat Biotechnol. 2019;37(8):852–857. doi:10.1038/s41587-019-0209-9

17. Callahan BJ, McMurdie PJ, Rosen MJ, Han AW, Johnson AJA, Holmes SP. DADA2: High-resolution sample inference from Illumina amplicon data. Nat Methods. 2016;13(7):581–583. doi:10.1038/nmeth.3869

18. Rognes T, Flouri T, Nichols B, Quince C, Mahé F. VSEARCH: a versatile open source tool for metagenomics. PeerJ. 2016;4:e2584. doi:10.7717/peerj.2584

19. Quast C, Pruesse E, Yilmaz P, et al. The SILVA ribosomal RNA gene database project: improved data processing and web-based tools. Nucleic Acids Res. 2013;41(Database issue):D590–596. doi:10.1093/nar/gks1219

20. Abarenkov K, Zirk A, Piirmann T, et al. Full UNITE+INSD dataset for Fungi. Published online February 19, 2025. doi:10.15156/BIO/3301227

21. McMurdie PJ, Holmes S. phyloseq: an R package for reproducible interactive analysis and graphics of microbiome census data. PLoS One. 2013;8(4):e61217. doi:10.1371/journal.pone.0061217

22. Davis NM, Proctor DM, Holmes SP, Relman DA, Callahan BJ. Simple statistical identification and removal of contaminant sequences in marker-gene and metagenomics data. Microbiome. 2018;6(1):226. doi:10.1186/s40168-018-0605-2

23. Oksanen J, Simpson GL, Blanchet FG, et al. vegan: Community Ecology Package. Published online September 6, 2001:2.7–3. doi:10.32614/CRAN.package.vegan

24. Kalan L, Loesche M, Hodkinson BP, et al. Redefining the Chronic-Wound Microbiome: Fungal Communities Are Prevalent, Dynamic, and Associated with Delayed Healing. mBio. 2016;7(5):e01058–16. doi:10.1128/mBio.01058-16

25. Costello EK, Lauber CL, Hamady M, Fierer N, Gordon JI, Knight R. Bacterial community variation in human body habitats across space and time. Science. 2009;326(5960):1694–1697. doi:10.1126/science.1177486

26. Kim JH, Son SM, Park H, et al. Taxonomic profiling of skin microbiome and correlation with clinical skin parameters in healthy Koreans. Sci Rep. 2021;11(1):16269. doi:10.1038/s41598-021-95734-9

27. Townsend EC, Xu K, De La Cruz K, et al. Still not sterile: viability-based assessment of the skin microbiome following pre-surgical application of a broad-spectrum antiseptic reveals transient pathogen enrichment and long-term recovery. Microbiol Spectr. 2025;13(5):e0287324. doi:10.1128/spectrum.02873-24

28. Cassir N, Papazian L, Fournier PE, Raoult D, La Scola B. Insights into bacterial colonization of intensive care patients’ skin: the effect of chlorhexidine daily bathing. Eur J Clin Microbiol Infect Dis. 2015;34(5):999–1004. doi:10.1007/s10096-015-2316-y

29. Popovich KJ, Lyles R, Hayes R, et al. Relationship between chlorhexidine gluconate skin concentration and microbial density on the skin of critically ill patients bathed daily with chlorhexidine gluconate. Infect Control Hosp Epidemiol. 2012;33(9):889–896. doi:10.1086/667371

30. Vernon MO, Hayden MK, Trick WE, et al. Chlorhexidine gluconate to cleanse patients in a medical intensive care unit: the effectiveness of source control to reduce the bioburden of vancomycin-resistant enterococci. Arch Intern Med. 2006;166(3):306–312. doi:10.1001/archinte.166.3.306

31. Sebbane N, Abramovitz I, Kot-Limon N, Steinberg D. Mechanistic Insight into the Anti-Bacterial/Anti-Biofilm Effects of Low Chlorhexidine Concentrations on Enterococcus faecalis-In Vitro Study. Microorganisms. 2024;12(11):2297. doi:10.3390/microorganisms12112297

32. Mougeot JLC, Beckman MF, Bahrani Mougeot F, Horton JM. Cutaneous Microbiome Profiles Following Chlorhexidine Treatment in a 72-Hour Daily Follow-Up Paired Design: a Pilot Study. Microbiol Spectr. 2022;10(3):e0175321. doi:10.1128/spectrum.01753-21

33. Kates AE, Zimbric ML, Mitchell K, Skarlupka J, Safdar N. The impact of chlorhexidine gluconate on the skin microbiota of children and adults: A pilot study. Am J Infect Control. 2019;47(8):1014–1016. doi:10.1016/j.ajic.2019.01.024

34. Wiemken TL, Ericsson AC. Chlorhexidine gluconate does not result in epidermal microbiota dysbiosis in healthy adults. Am J Infect Control. 2021;49(6):769–774. doi:10.1016/j.ajic.2020.11.021

35. Ray Mohapatra A, Jeevaratnam K. Inhibiting bacterial colonization on catheters: Antibacterial and antibiofilm activities of bacteriocins from Lactobacillus plantarum SJ33. J Glob Antimicrob Resist. 2019;19:85–92. doi:10.1016/j.jgar.2019.02.021

36. Kampf G. Acquired resistance to chlorhexidine - is it time to establish an “antiseptic stewardship” initiative? J Hosp Infect. 2016;94(3):213–227.

37. Hardy K, Sunnucks K, Gil H, et al. Increased Usage of Antiseptics Is Associated with Reduced Susceptibility in Clinical Isolates of Staphylococcus aureus. mBio. 2018;9(3):e00894–18. doi:10.1128/mBio.00894-18

38. Madden GR, Sifri CD. Antimicrobial Resistance to Agents Used for Staphylococcus aureus Decolonization: Is There a Reason for Concern? Curr Infect Dis Rep. 2018;20(8):26. doi:10.1007/s11908-018-0630-0

39. Horner C, Mawer D, Wilcox M. Reduced susceptibility to chlorhexidine in staphylococci: is it increasing and does it matter? J Antimicrob Chemother. 2012;67(11):2547–2559. doi:10.1093/jac/dks284

40. Zhang E, Tanaka T, Tsuboi R, Makimura K, Nishikawa A, Sugita T. Characterization of Malassezia microbiota in the human external auditory canal and on the sole of the foot. Microbiol Immunol. 2012;56(4):238–244. doi:10.1111/j.1348-0421.2012.00430.x

41. Nascimento T, Inácio J, Guerreiro D, et al. Can chlorhexidine gluconate baths reduce fungal colonisation in intensive care unit patients? Antimicrob Resist Infect Control. 2025;14(1):87. doi:10.1186/s13756-025-01606-6

42. Fathilah AR, Himratul-Aznita WH, Fatheen ARN, Suriani KR. The antifungal properties of chlorhexidine digluconate and cetylpyrinidinium chloride on oral Candida. J Dent. 2012;40(7):609–615. doi:10.1016/j.jdent.2012.04.003

43. Abdolrasouli A, Armstrong-James D, Ryan L, Schelenz S. In vitro efficacy of disinfectants utilised for skin decolonisation and environmental decontamination during a hospital outbreak with Candida auris. Mycoses. 2017;60(11):758–763. doi:10.1111/myc.12699

44. Frost SA, Alogso MC, Metcalfe L, et al. Chlorhexidine bathing and health care-associated infections among adult intensive care patients: a systematic review and meta-analysis. CritCare. 2016;20(1):379. doi:10.1186/s13054-016-1553-5

45. Giri VK, Kegerreis KG, Ren Y, et al. Chlorhexidine Gluconate Bathing Reduces the Incidence of Bloodstream Infections in Adults Undergoing Inpatient Hematopoietic Cell Transplantation. Transplant Cell Ther. 2021;27(3):262.e1-262.e11. doi:10.1016/j.jtct.2021.01.004

46. Artese AL, Sainvil M, Fish LJ, et al. Exploring facilitators and barriers to daily chlorhexidine gluconate bathing in adult patients undergoing hematopoietic stem cell transplantation. Support Care Cancer. 2024;32(12):833. doi:10.1007/s00520-024-09037-6

47. Kalan LR, Meisel JS, Loesche MA, et al. Strain- and Species-Level Variation in the Microbiome of Diabetic Wounds Is Associated with Clinical Outcomes and Therapeutic Efficacy. Cell Host Microbe. 2019;25(5):641–655.e5. doi:10.1016/j.chom.2019.03.006

48. Loesche M, Gardner SE, Kalan L, et al. Temporal Stability in Chronic Wound Microbiota Is Associated With Poor Healing. J Invest Dermatol. 2017;137(1):237–244. doi:10.1016/j.jid.2016.08.009

49. Campbell AE, McCready-Vangi AR, Uberoi A, et al. Variable staphyloxanthin production by Staphylococcus aureus drives strain-dependent effects on diabetic wound-healing outcomes. Cell Rep. 2023;42(10):113281. doi:10.1016/j.celrep.2023.113281

