## Supplemental Figures for "Long-Term Daily Chlorhexidine Foot Cleansing Reduces Staphylococcal Burden on the Feet of People with Prior Diabetic Foot Complications"

Supplemental Table 1: Self-reported Foot Care of Randomized Participants at Baseline by Treatment Group

| | **Chlorhexidine Group (n=47)** | **Soap and Water Group (n=40)** | **Total (n=87)** | **P value** |
|---|---|---|---|---|
| In the past 4 weeks, how often have you… | | | | |
| Washed your feet with soap and water | | | | 0.76 |
| Not at all | 0 (0) | 0 (0) | 0 (0) | |
| About once or twice each month | 1 (3) | 0 (0) | 1 (1) | |
| Once each week | 3 (8) | 2 (6) | 5 (7) | |
| Several times each week | 13 (34) | 8 (26) | 21 (30) | |
| Daily | 21 (55) | 21 (68) | 42 (61) | |
| Washed your feet with a washcloth | | | | 0.28 |
| Not at all | 6 (16) | 1 (3) | 7 (10) | |
| About once or twice each month | 0 (0) | 2 (6) | 2 (3) | |
| Once each week | 3 (8) | 2 (6) | 5 (7) | |
| Several times each week | 10 (26) | 8 (26) | 18 (26) | |
| Daily | 19 (50) | 18 (58) | 37 (54) | |
| Used lotion on your feet | | | | 0.93 |
| Not at all | 12 (26) | 12 (30) | 24 (28) | |
| About once or twice each month | 1 (2) | 1 (3) | 2 (2) | |
| Once each week | 6 (13) | 5 (13) | 11 (13) | |
| Several times each week | 9 (19) | 5 (13) | 14 (16) | |
| Daily | 19 (40 | 17 (43) | 36 (41) | |

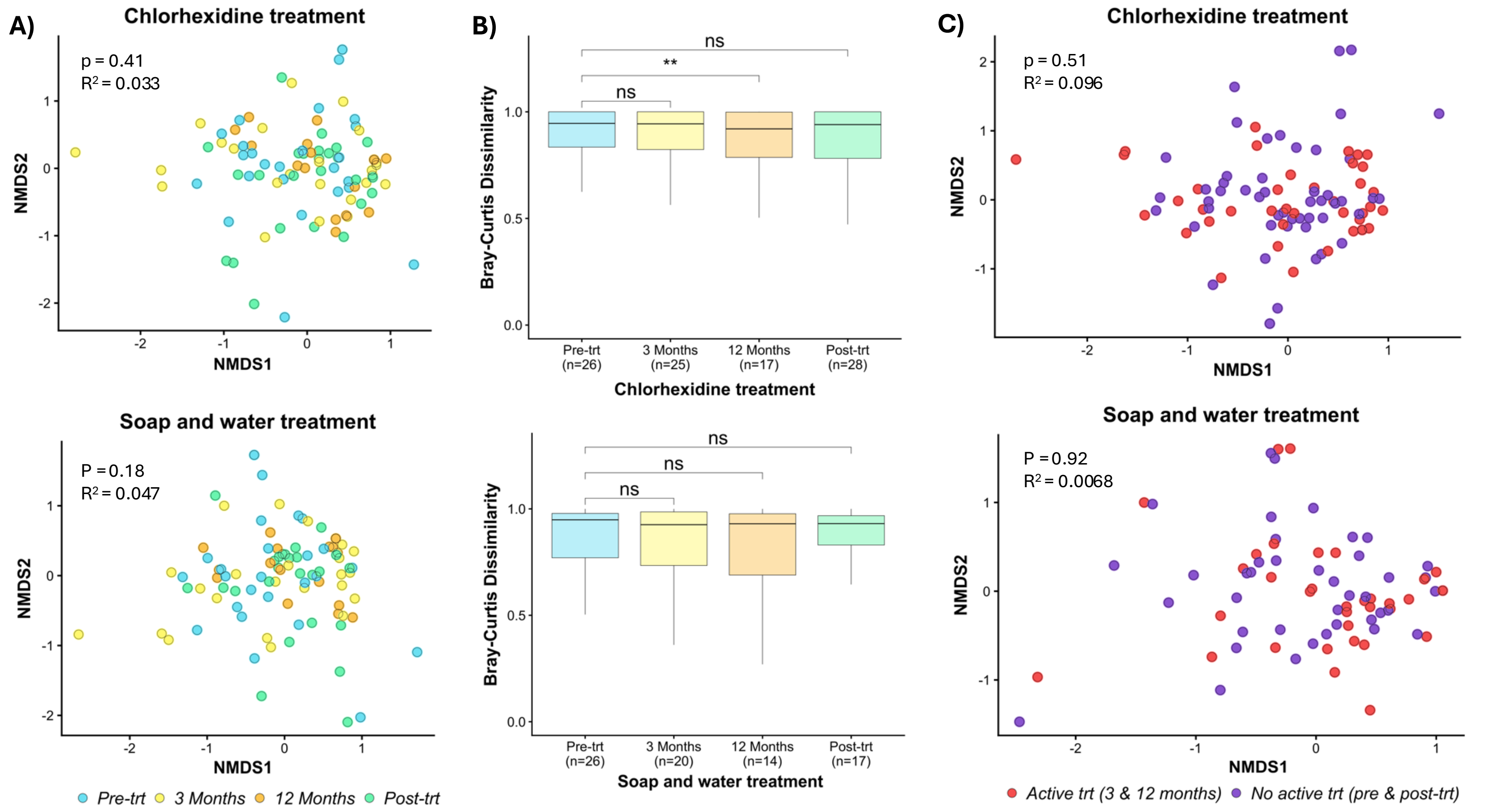


**Supplemental Figure 1: Chlorhexidine treatment caused no significant changes in the fungal microbiome.** For fungal diversity analysis, samples with low counts were removed, OTU counts were normalized with rarefaction, and data were merged to the genus level. Samples were then separated by visit for both chlorhexidine (pre-trt n = 26, 3 months n = 25, 12 months n = 17, post-trt n = 28) and soap-and-water (pre-trt n = 26, 3 months n = 20, 12 months n = 14, post-trt n = 17) treatments. (A) Differences in the relative abundances of genera in each sample were measured using Bray–Curtis dissimilarity and visualized on an NMDS plot. (B) The average distance of all samples from pre-treatment samples was measured and separated by visit to observe whether distance increased with treatment. (C) Bray–Curtis distances between samples were visualized on an NMDS plot. Samples were grouped by whether there was active treatment (3-month and 12-month) shown in red, or no active treatment (pre-treatment and post-treatment) shown in purple. Statistically significant differences between groups were calculated using PERMANOVA, with Benjamini–Hochberg p-value adjustment. Statistically significant differences between groups for pairwise comparisons were calculated using a Wilcoxon rank-sum test, with Benjamini–Hochberg p-value adjustment. * $p < 0.05$, ** $p < 0.01$, *** $p < 0.001$, **** $p < 0.0001$.

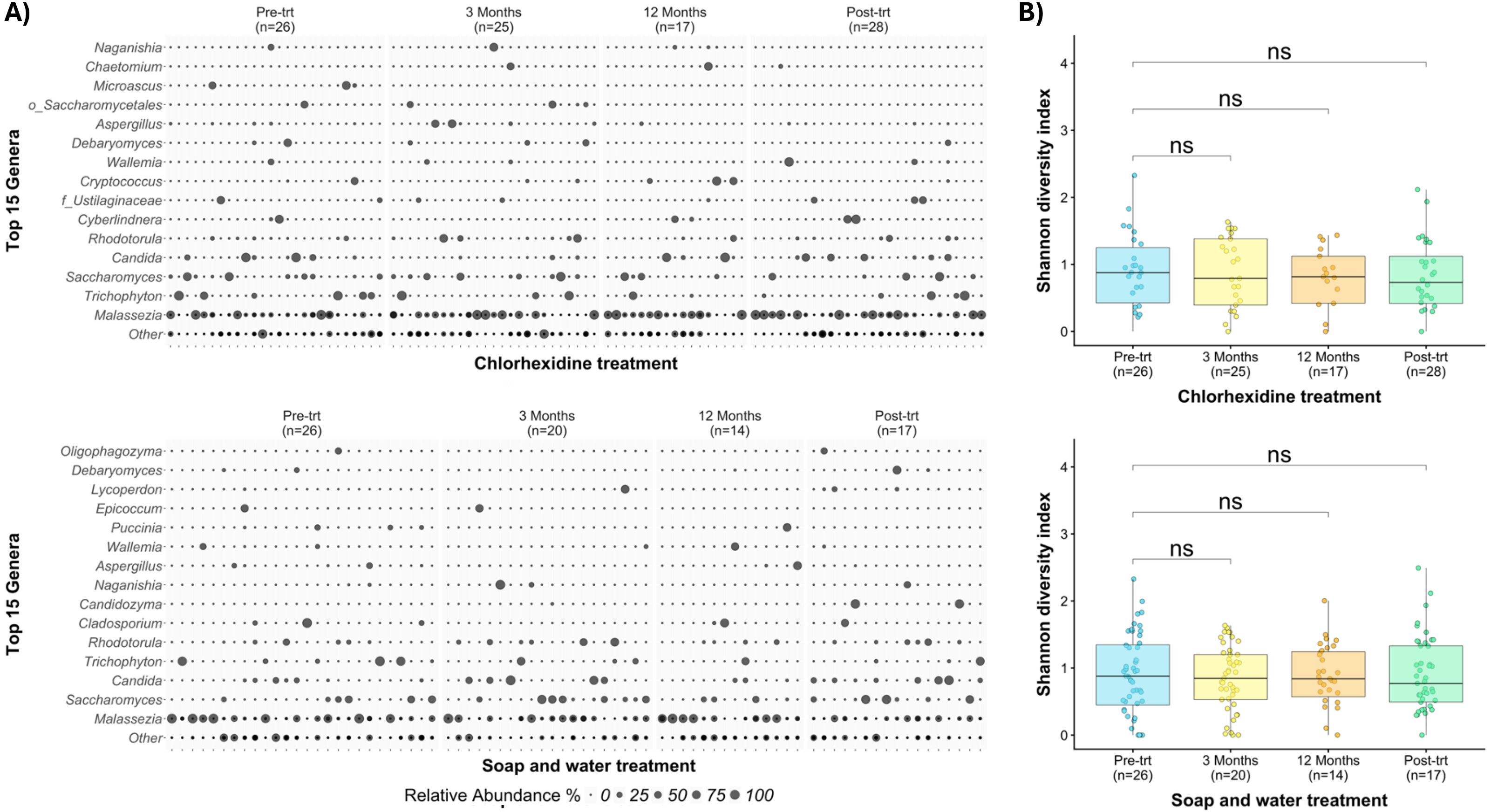


**Supplemental Figure 2: Chlorhexidine treatment does not cause significant changes in the relative abundance and Shannon diversity of fungal genera.** (A) The relative abundances of the top 15 fungal genera were compared separately over the course of treatment with chlorhexidine (pre-trt n = 26, 3 months n = 25, 12 months n = 17, post-trt n = 28) and soap-and-water (pre-trt n = 26, 3 months n = 20, 12 months n = 14, post-trt n = 17). (B) For fungal diversity analysis, samples with low counts were removed, OTU counts were normalized with rarefaction, and data were merged to the genus level. Shannon diversity was measured in each sample and compared separately over the course of each treatment. Statistically significant differences in averages between pre-treatment samples and subsequent visits were calculated using a Wilcoxon rank-sum test, with Benjamini–Hochberg p-value adjustment. * $p < 0.05$, ** $p < 0.01$, *** $p < 0.001$, **** $p < 0.0001$.

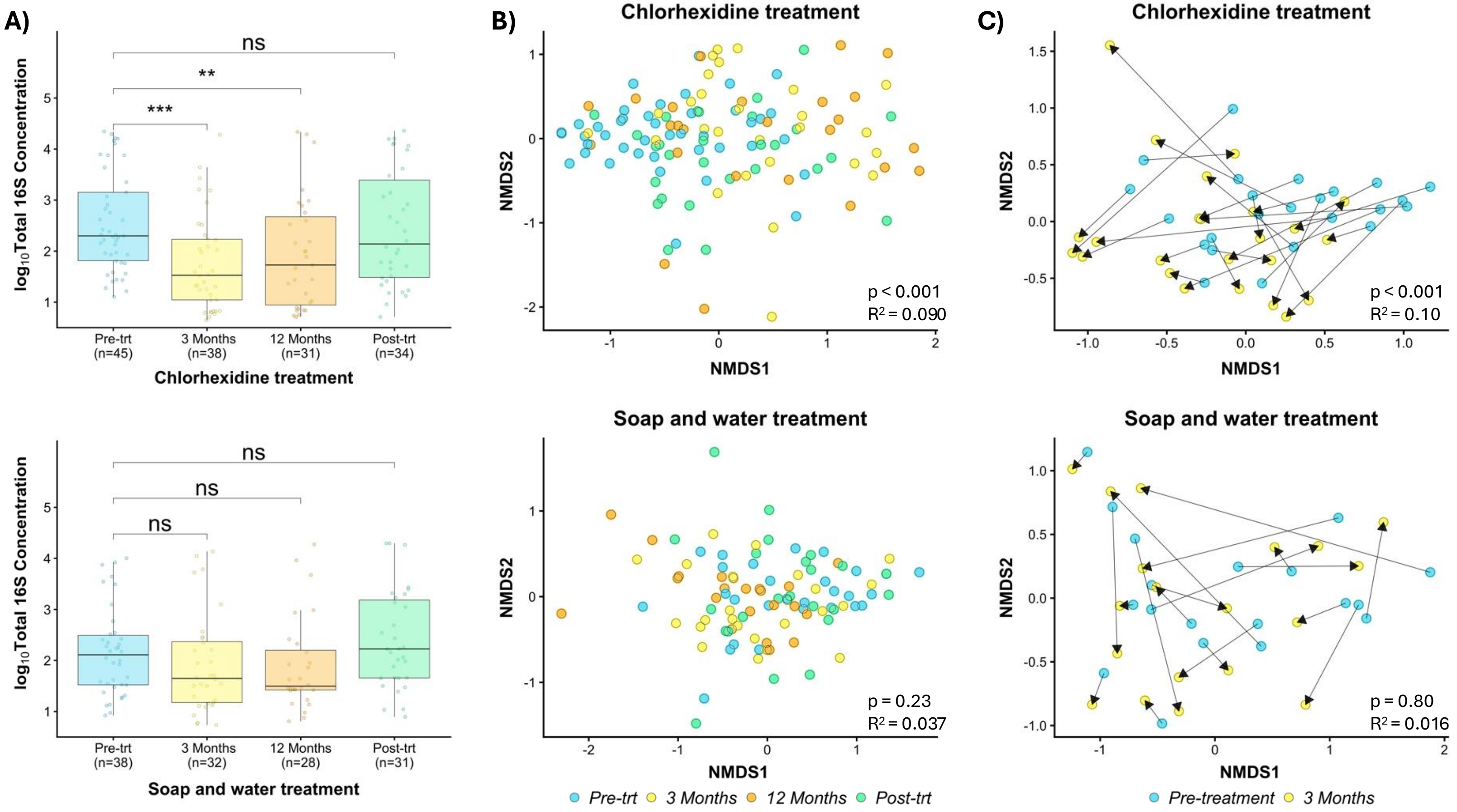


**Supplemental Figure 3: Chlorhexidine treatment causes significant decreases in bioburden and consistent directional shift in the core microbiome.** (A)The log concentrations of total 16S in each sample were measured using dPCR and compared separately over the course of treatment for chlorohexidine (pre-trt n= 45 months n =38, 12 months n=31, post-trt n=34) and soap-and-water (pre-trt n= 38 months n =32, 12 months n=21, post-trt n=31). (B) For bacterial diversity analysis, samples with few counts were removed, OTU counts were normalized with rarefaction and merged to genus level. Samples were then separated by visit for both chlorohexidine (pre-trt n= 42 months n =27, 12 months n=25, post-trt n=28) and soap-and-water (pre-trt n= 27, months n=24, 12 months n=22, post-trt n=24) treatments. The differences in the relative abundances of genera in each sample were measured using Bray-Curtis dissimilarity and visualized on an NMDS plot. (C) To observe changes in an individual, Bray-Curtis distances between samples from pre-treatment and after 12 months of treatment were visualized on an NMDS plot lines connecting samples from the same patient. Arrows indicate the direction of shift from pre-treatment to after 12 months of treatment for an individual. Statically significant differences between groups were calculated using PERMANOVA, with Benjamini-Hochberg p-value adjustment. Statically significant differences between groups for pairwise comparisons were calculated using a Wilcoxon-rank sum test, with Benjamini-Hochberg p-value adjustment. * p<0.05, ** p<0.01, *** p<0.001, **** p<0.0001.

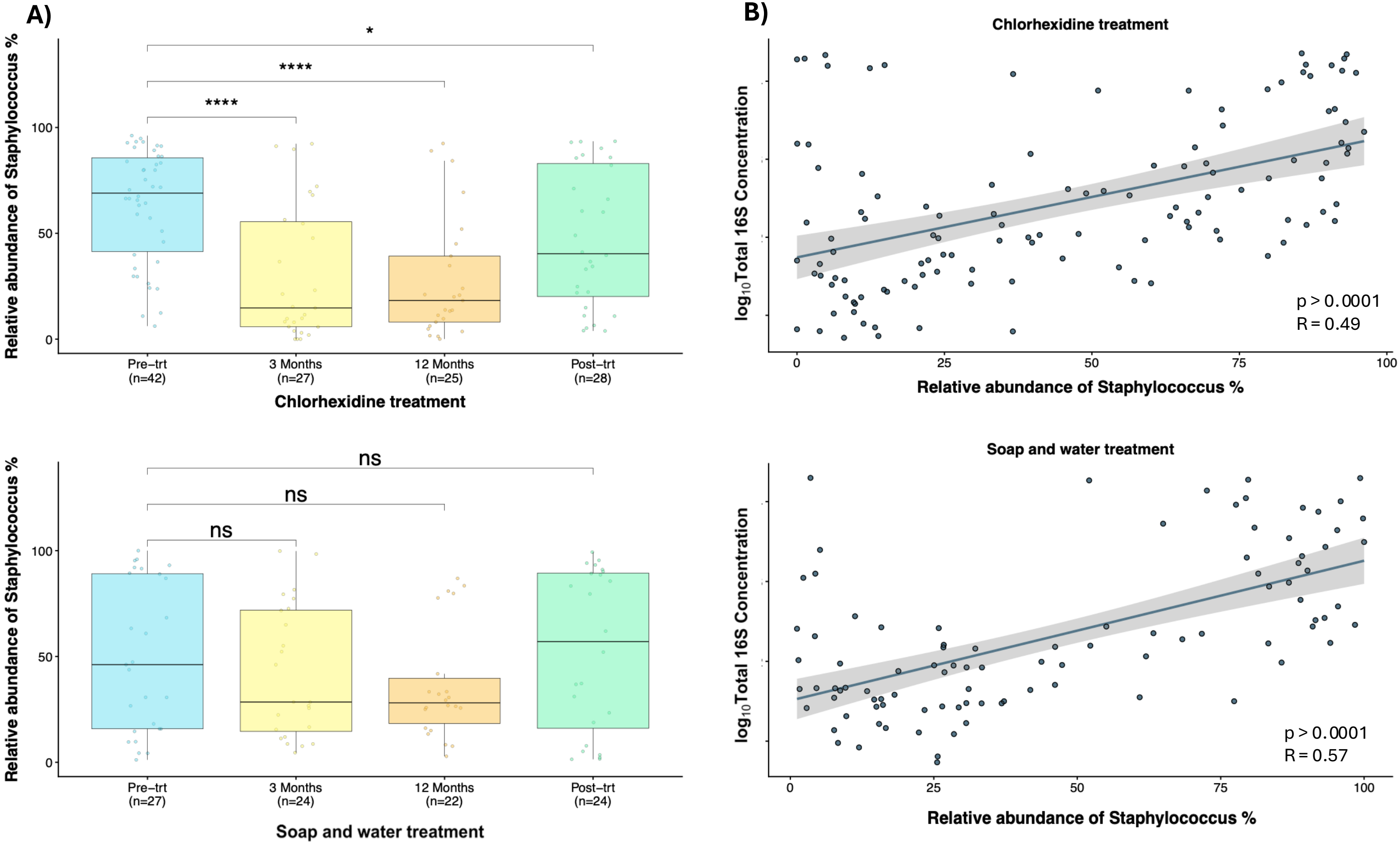


**Supplemental Figure 4: Chlorhexidine treatment causes significant decreases in staphylococcus abundance that is correlated with a decrease in bioburden.** (A) The relative abundances of *Staphylococcus* in each sample, determined with 16S sequencing, were compared separately over the course of treatment with chlorohexidine (pre-trt n= 26 months n =25, 12 months n=17, post-trt n=28) and soap-and-water (pre-trt n= 26, months n=20, 12 months n=14, post-trt n=17). Statically significant differences in averages between pre-treatment samples and the following visits were calculated using a Wilcoxon-rank sum test, with Benjamini-Hochberg p-value adjustment. * p<0.05, ** p<0.01, *** p<0.001, **** p<0.0001. (B) The log concentrations of total 16S were measured using dPCR and compared to the relative abundances of *Staphylococcus* to observe if changes were proportional to each other. A significant linear correlation was measured using Spearman rank correlations.
